# A multivariable Mendelian randomization analysis disentangling the causal relations between abdominal obesity, non-alcoholic fatty liver disease and cardiometabolic diseases

**DOI:** 10.1101/2021.09.28.21264201

**Authors:** William Pelletier, Éloi Gagnon, Émilie Gobeil, Jérôme Bourgault, Hasanga D. Manikpurage, Ina Maltais-Payette, Erik Abner, Nele Taba, Tõnu Esko, Patricia L. Mitchell, Nooshin Ghodsian, Jean-Pierre Després, Marie-Claude Vohl, André Tchernof, Sébastien Thériault, Benoit J. Arsenault

## Abstract

**Background:** Observational studies have linked obesity and especially abdominal obesity to non-alcoholic fatty liver disease (NAFLD). These traits are also associated with type 2 diabetes (T2D) and coronary artery disease (CAD) but the causal factor(s) underlying these associations remain unexplored.

**Methods:** We used a multivariable Mendelian randomization (MVMR) study design to determine whether obesity (defined using body mass index [BMI]) and abdominal obesity (defined using waist circumference) were causally associated with NAFLD using publicly available genome-wide association study (GWAS) summary statistics of the UK Biobank (n>450,000) and a GWAS meta-analysis of NAFLD (8434 cases and 770,180 control). A MVMR study design was also used to determine the respective causal contributions of waist circumference and NAFLD to T2D and CAD using additional GWAS summary statistics of the DIAGRAM (74,124 cases and 824,006 controls) and CARDIoGRAMplusC4D (122,733 cases and 424,528 controls) consortia.

**Results:** In univariable Mendelian randomization analyses, both BMI and waist circumference were associated with NAFLD. NAFLD was not associated with obesity or abdominal obesity. In MVMR analyses, waist circumference was associated with NAFLD when accounting for BMI (OR per 1-standard deviation increase = 2.56 95% CI: 1.39-4.69, p=2.4e-03) and BMI was not associated with NAFLD when accounting for waist circumference (0.81 95% CI: 0.5-1.31, p =3.9e-01). In MVMR analyses accounting for NAFLD, waist circumference remained strongly associated with both T2D (3.25 95% CI: 2.87-3.68, p=5.1e-77) and CAD (1.62 95% CI: 1.48-1.76, p=6.5e-28).

**Conclusions:** These results identified abdominal obesity as a strong, independent and causal contributor to NAFLD, T2D and CAD, suggesting that interventions targeting abdominal obesity rather than body weight per se should be prioritized for the prevention and management of cardiometabolic diseases.

## Introduction

Non-alcoholic fatty liver disease (NAFLD) is characterized by hepatic lipid accumulation ranging from simple steatosis (>5% of liver weight is lipids) to non-alcoholic steatohepatitis (NASH, presence of inflammation) (Chalasani et al., 2018). Although simple steatosis is relatively benign, more severe forms of NAFLD such as NASH and hepatic fibrosis can lead to liver cirrhosis and hepatocellular carcinoma. Approximately 25% of the adult population globally is affected by NAFLD with the prevalence rapidly increasing and potentially becoming the leading cause of liver failure in the United States by 2025 (Charlton et al., 2011; Younossi et al., 2016). Obesity and body fat distribution are closely linked with NAFLD (Ross et al., 2020). In the INSPIRE ME study, a large international imaging study by computed tomography, waist circumference was closely associated with liver fat accumulation independently of body mass index (BMI) (Nazare et al., 2015).

Studies have also shown that both liver fat accumulation/NAFLD and waist circumference are associated with CAD and T2D (Fabbrini et al., 2009; Kotronen & Yki-Järvinen, 2008; Ndumele et al., 2011; Tilg, Moschen, & Roden, 2017). However, whether or not these relationships are causal remains to be elucidated and, more importantly, whether or not agents aimed at targeting NAFLD will ultimately decrease the risk of either T2D or CAD is unknown. In a previous investigation, we showed a strong genetic correlation between NAFLD, waist circumference, T2D and CAD (Ghodsian, 2021). However, little is known about the directionality of these relationships and whether NAFLD lies in the causal pathway linking abdominal obesity and T2D/CAD.

In order to gain insight about the causality and directionality of these associations, new causal inference methods such as Mendelian randomization (MR) have been developed. MR uses genetic variants (which are randomly distributed at meiosis) such as single-nucleotide polymorphisms (SNPs), as instruments to infer causality. This method is comparable to a randomized control trial in which participants are naturally randomized based on the presence or absence of genetic variants that influence traits of interest. In a previous MR study, waist-to-hip ratio (WHR) adjusted for BMI was strongly associated with T2D and CAD (Emdin et al., 2017). However, we do not know if similar associations exist for NAFLD.

Extensions of the MR design, such as bidirectional MR and multivariable MR (MVMR), help in clarifying causal relations. Bidirectional MR refers to an analysis where both traits are alternately evaluated as exposure and outcome. This method has the potential to remove reverse causation bias by asserting the directionality of the relationship (Welsh et al., 2010). Multivariable MR is used when multiple genetic variants are associated to two or more exposures. It conditions the effects of the SNPs of each exposure together to assess the effect of each exposure independently on the outcome. This method allows the identification of the true causal factor when two exposures share genetic variants as if they had been adjusted for one another (Sanderson, Davey Smith, Windmeijer, & Bowden, 2019).

Here, we first used bidirectional and multivariable MR designs to investigate the respective causal contributions of obesity (defined using BMI) and abdominal obesity (defined using waist circumference) to NAFLD. Second, using a similar strategy, we aimed to determine if abdominal obesity and NAFLD are independent causal risk factors for T2D and CAD.

## Results

### Bi-directional associations between obesity and NAFLD

The study design and cohorts used in this MR study are presented in Figure 1. We first investigated the bi-directional associations between obesity (defined by BMI or waist circumference) and NAFLD using Inverse Variance Weighted (IVW)-MR and other robust analyses described in the Methods section. Results from all MR methods (Figure 2) suggest that BMI and waist circumference are both causally associated with NAFLD, while NAFLD is not associated with obesity or abdominal obesity. A one standard deviation (SD)-higher waist circumference had an odds ratio (OR) of 2.03 (95% confidence interval [CI]: 1.76-2.34, p=6.7e-23) for NAFLD and a one SD-higher BMI had an OR of 1.67 (95 % CI: 1.49-1.87, p=1.0e-18) (Figure 3). Specifically, every additional centimeter of waist circumference increased the risk of NAFLD (OR=1.05, CI: 1.04-1.07, p=6.7e-23) and every BMI point increased the risk of NAFLD (OR=1.11, CI: 1.09-1.14, p=1.0e-18). When BMI and waist circumference were assessed together in MVMR, only waist circumference (OR=2.56 95% CI: 1.39-4.69, p=2.4e-03) retained a robust association with NAFLD, while the effect of BMI was not significant (OR=0.81 95% CI=0.50-1.31, p=3.9e-01) (Figure 3). Similar results were obtained using the Genetic Investigation of Anthropometric Traits (GIANT) consortium as study exposures for WC and BMI (Supplementary Figures 1-2).

**Figure 1.**
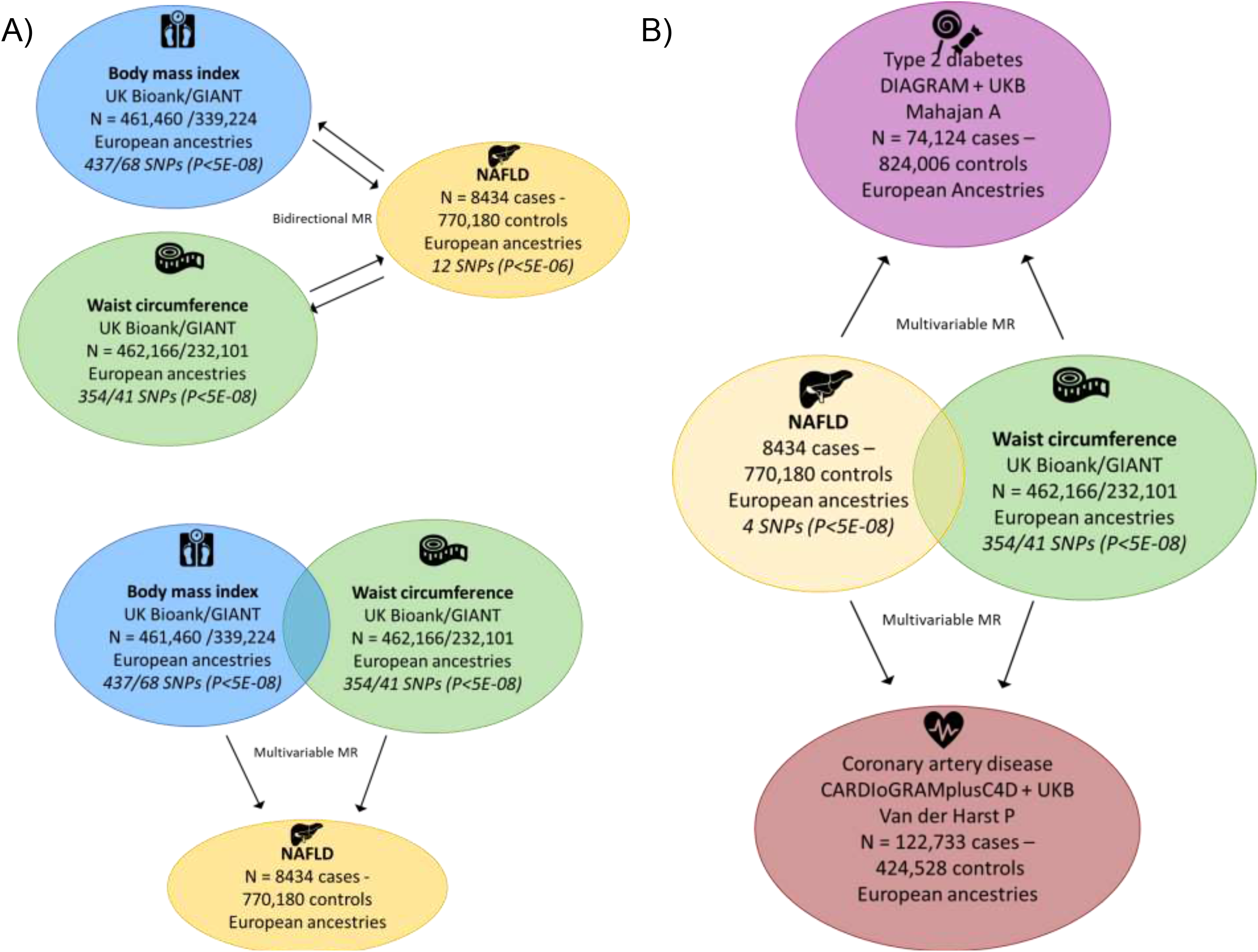
Schematic overview of the analytical framework used to disentangle the causal relationships between abdominal obesity, non-alcoholic fatty liver disease and type 2 diabetes. A) Design of the bi-directional associations between obesity (assessed using the body mass index and waist circumference) and non-alcoholic fatty liver disease (NAFLD) using univariable and multivariable mendelian randomization (MVMR). B) Design of the MVMR analysis investigating the respective contributions of abdominal obesity and NAFLD on type 2 diabetes and coronary artery disease.

**Figure 2.**
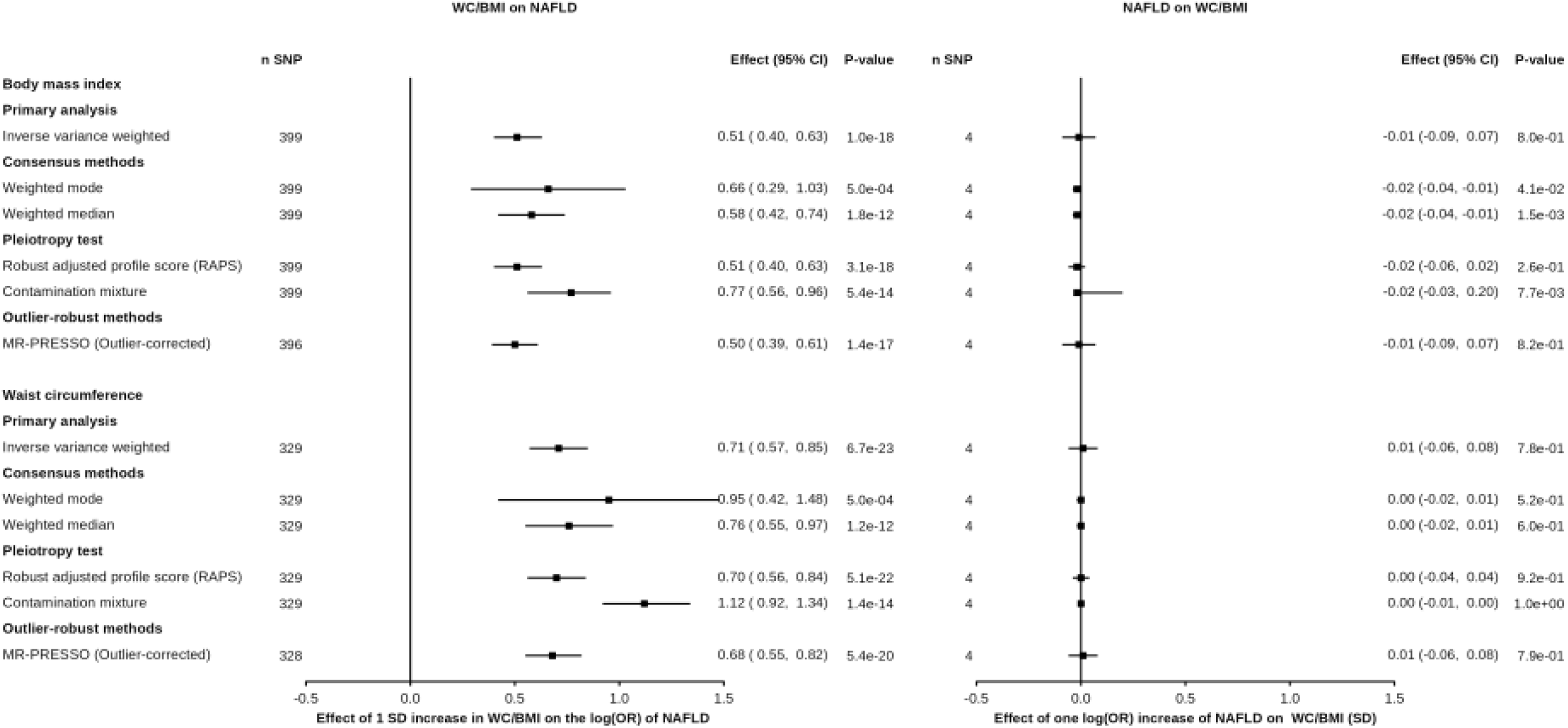
Bi-directional associations between obesity and non-alcoholic fatty liver disease (NAFLD). A) association of body mass index and waist circumference (exposures) with NAFLD (outcome) using inverse-variance weighted Mendelian randomization (IVW-MR) and robust MR analyses B) association of NAFLD (exposure) with body mass index and waist circumference (outcomes) using IVW-MR and robust MR analyses.

**Figure 3.**
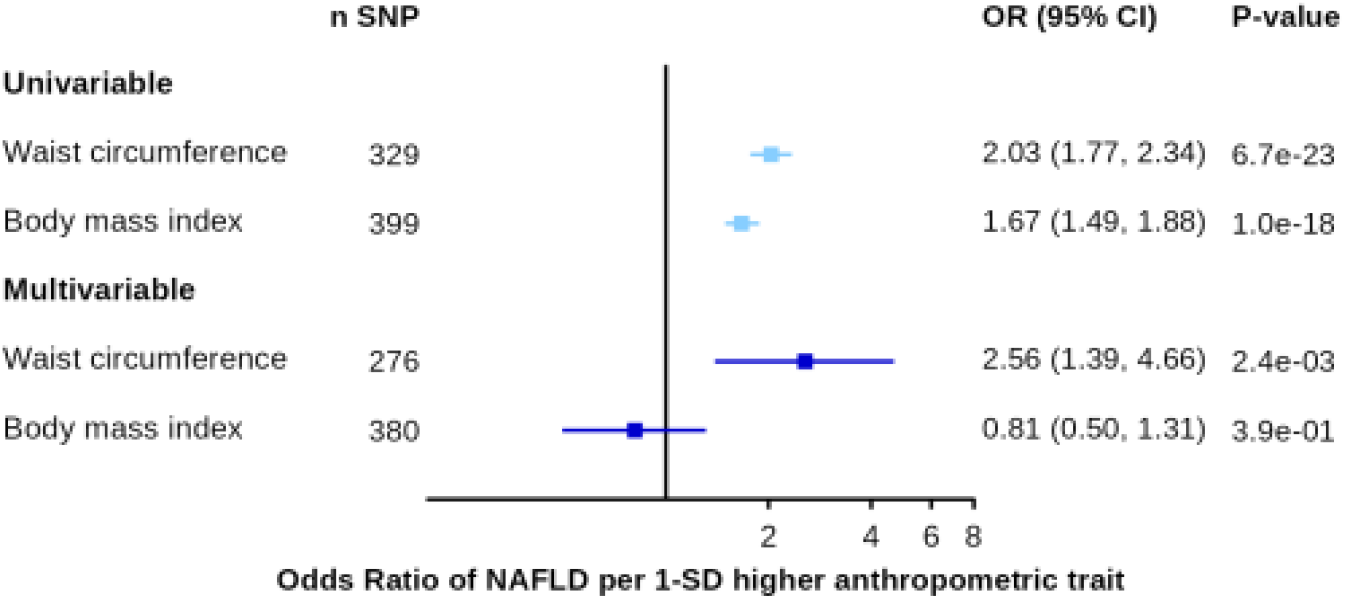
Association between waist circumference and body mass index with non-alcoholic fatty liver disease using univariable and multivariable Mendelian randomization.

We investigated the association of WHR adjusted for BMI with NAFLD using multiple MR methods. A higher genetically predicted WHR adjusted for BMI was associated with NAFLD across all MR methods (Figure 4). Altogether, these analyses provide evidence that body fat distribution patterns consistent with higher visceral fat accumulation is an important determinant of NAFLD, regardless of subcutaneous fat accumulation or obesity indices that do not take into account body composition such as the BMI.

**Figure 4.**
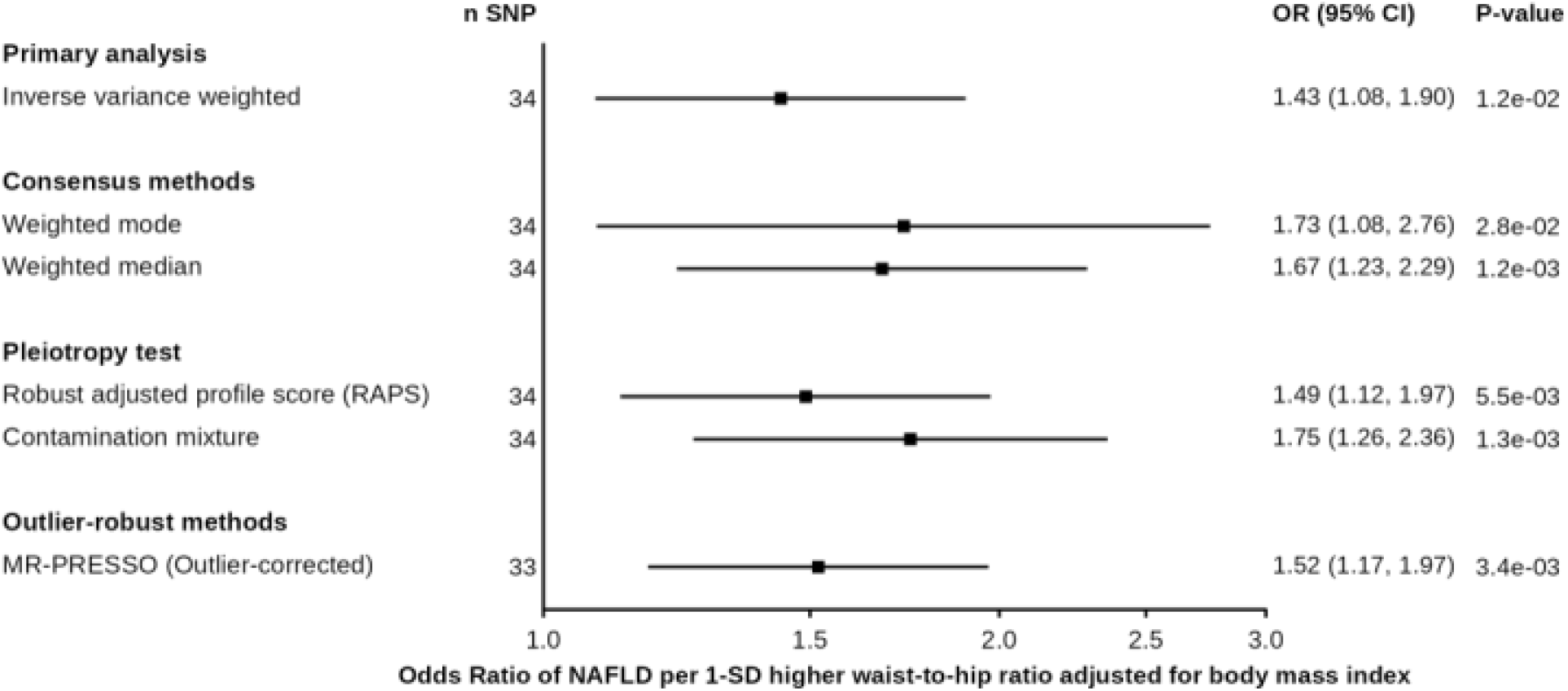
Association between waist-to-hip ratio adjusted for body mass index and non-alcoholic fatty liver disease using multiple Mendelian randomization methods.

### Contributions of abdominal obesity and non-alcoholic fatty liver disease to type 2 diabetes and CAD

In IVW-MR, a 1-SD increment in waist circumference or 1-log(OR) increment of NAFLD was associated with T2D (OR=3.68 95% CI: 3.29-4.13, p=2.9e-110 and OR=1.24 95% CI: 1.05-1.47, p=1.2e-02, respectively for waist circumference and NAFLD) (Figure 5, left panel). NAFLD MR estimates on T2D were consistent for all MR methods and significant for most MR methods (Supplementary figure 3). When waist circumference and NAFLD were assessed together in MVMR, waist circumference (OR=3.25 95% CI: 2.87-3.68, p=5.1e-77 and NAFLD (1.19 95% CI: 1.12-1.27, p=1.6e-07) increased the risk of T2D. When deriving WC instruments from GIANT, NAFLD was not associated with T2D when accounting for WC in MVMR (OR=1.05 95% CI: 0.72-1.53, p=8.0e-01) (Supplementary Figures 4 left panel). This inconsistency between study cohorts is likely a result of the minimal genetic coverage in GIANT. GIANT summary statistics only included one of the four genetic instruments of NAFLD or their LD proxy R2>0.8.

**Figure 5.**
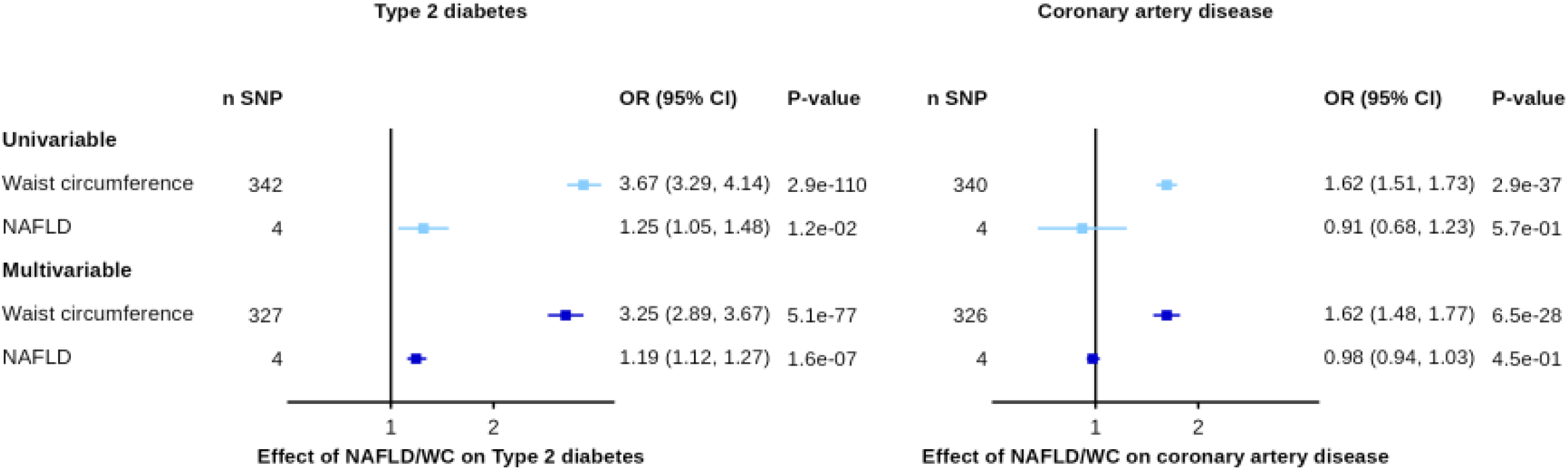
Association between waist circumference and coronary artery disease and type 2 diabetes before and after accounting for non-alcoholic fatty liver disease using univariable and multivariable Mendelian randomization.

In IVW-MR, a 1-SD increment in waist circumference was associated with CAD (OR=1.62 95% CI: 1.50-1.74, p=2.9e-37), while NALFD was not associated with CAD (Figure 5, right panel). When waist circumference and NAFLD were assessed together in MVMR, waist circumference retained a robust association with CAD (OR=1.62 95% CI: 1.48-1.76, p=6.5e-28) while NAFLD was not associated with CAD. The results were similar when deriving WC instruments from GIANT (Supplementary Figure 4 right panel). Results of this analysis revealed that abdominal obesity is a stronger risk factor for T2D and CAD than NAFLD. Although abdominal obesity is closely associated with NAFLD, the impact of abdominal obesity on T2D and CAD is minimally mediated by NAFLD.

## Discussion

In this study, we explored the bi-directional relationships between total and abdominal obesity and NAFLD using MR. We found that total and abdominal obesity were causally linked to NAFLD while NAFLD was not causally linked to obesity. Results of our multivariable MR analysis suggest that waist circumference is causally linked to NAFLD regardless of body weight, while BMI is not causally linked with NAFLD once waist circumference is taken into account. Observational studies have documented similar associations (Jarvis et al., 2020). Liu et al. used bidirectional MR to explore the relationship between NAFLD, obesity, T2D and lipid traits (Liu et al., 2020). They found that both obesity and abdominal obesity had a causal effect on NAFLD. Our study provides additional support for a causal effect of abdominal obesity on NAFLD using a larger study sample size for the study outcome (NAFLD). Our study is, to our knowledge, the first to report using MVMR a causal association between abdominal obesity and NAFLD that is independent of body weight.

Having established a causal role of abdominal obesity on NAFLD and given the results of previous studies linking NAFLD to cardiometabolic diseases such as T2D and CAD, we explored whether NAFLD lies in the causal pathway linking abdominal obesity to T2D and CAD. In their MR investigation, Liu et al. also showed that NAFLD had a causal effect on T2D (OR: 1.3, 95% CI: 1.2, 1.4, p=8.3e-14) (Liu et al., 2020). They used 2 genetic instruments for NAFLD, making it impossible to perform robust MR analyses. In our analysis, NAFLD was similarly associated with T2D and robust MR analysis were consistent with a causal association. The association was slightly decreased when we accounted for abdominal obesity using MVMR, suggesting that part of the effect of NAFLD on T2D could be attributable to pleiotropic variants shared with abdominal fat accumulation. On the other hand, the point estimate of WC on T2D also only slightly decreased when we accounted for NAFLD, suggesting that the effect of WC on T2D is minimally mediated by NAFLD. Using MVMR, our results support, for the first time to our knowledge, that the association between abdominal obesity and T2D is independent of NAFLD. We also showed that the association between abdominal obesity and CAD is independent of NAFLD.

The inability of subcutaneous fat to expand by hyperplasia may partly explain why visceral fat accumulation occurs in genetically predisposed individuals (Tchernof & Després, 2013). These excess lipids are then stored in lean tissues such as the liver, heart and skeletal muscle promoting insulin resistance (Ross et al., 2020; Ye, Richard, Gévry, Tchernof, & Carpentier, 2021). The mechanisms by which visceral fat contributes to NAFLD may also possibly be explained by the “portal vein theory”(Rytka, Wueest, Schoenle, & Konrad, 2011). Visceral fat is mostly drained by the portal vein, which delivers its content to the liver and exposes it to high concentrations of free fatty acids and adipokines. These have been hypothesized to lead to metabolic changes in the liver which would ultimately lead to an increased production of VLDL particles, glucose and inflammatory mediators as well as decreased insulin extraction, potentially leading to T2D and atherosclerosis.

From a clinical perspective, results of this study support the idea that previously reported associations between an elevated BMI and NAFLD may be explained by preferential abdominal fat accumulation reflected by higher waist circumference. Indeed, a significant number of individuals with elevated BMI have excess visceral fat increasing their risk of NAFLD (Liu et al., 2020; Loomis et al., 2016; Miyake et al., 2013; Wang, Guo, & Lu, 2016). Our results also underline the limitations of the sole use of BMI in clinical practice to assess the risk associated with obesity/ectopic fat distribution. The failure of BMI to capture cardiometabolic risk had already been suggested by observation and MR studies (Nazare et al., 2015; Ross et al., 2020; Snijder, van Dam, Visser, & Seidell, 2006). Our study adds evidence supporting waist circumference as a simple tool to assess obesity-related health hazards.

These results should encourage clinical interventions focused on visceral fat reduction, not only overall body weight reduction, to prevent cardiometabolic diseases such as NAFLD, T2D and CAD. Visceral fat can be targeted with physical activity and dietary interventions even in the absence of weight loss. A weight loss of about 5% can result in a 15–25% visceral fat reduction (I. J. Neeland et al., 2019). The Mediterranean diet as well as diets lower in fat and/or carbohydrate may be effective ways of reducing visceral fat, especially in physically active individuals (Gepner et al., 2018; Ross et al., 2020; Verheggen et al., 2016). There is also evidence that thiazolidinediones (TZDs) such as pioglitazone and rosiglitazone, used in the treatment of T2D, increase subcutaneous adipocytes’ storage capacity and lower T2D risk (Unger, 2008). Results of the VICTORY trial, a study aimed at assessing the safety and efficiency of rosiglitazone on saphenous vein graft atherosclerosis and the cardiometabolic risk profile, showed that rosiglitazone treatment induced a 3 kg weight gain over 12 months and no change in visceral adiposity (Bertrand et al., 2010). Pioglitazone has also been shown to reduce hepatic steatosis and inflammation in patients with NASH (Sanyal et al., 2010) thereby providing randomized clinical trial support to our MR findings. Semaglutide, a glucagon-like protein-1 (GLP-1) receptor agonist, has recently been shown to increase the rate of NASH resolution compared with placebo (Newsome et al., 2020). A recent study on another GLP-1 receptor agonist liraglutide also recently provided evidence that this class of drug may induce a preferential loss in visceral adipose tissue (Ian J. Neeland et al.).

An important strength of the current study is the use of the largest NAFLD dataset available to date. Additionally, the use of a MVMR design that is not subject to confounding factors and reverse causality bias enables the estimation of the direct effect of closely related risk factors on cardiometabolic outcomes. Our study, however, has limitations. Few genome wide significant NAFLD SNPs were uncovered even though the largest GWAS was used. This could introduce winner’s curse bias mainly for SNPs close to the genome-wide significance p-value threshold. The winners’ curse may decrease the robustness of these genetic instruments and could bias the effect of NAFLD on cardiometabolic traits toward the null. However, the low number of genome-wide genetic instruments should have little impact on the analysis using NAFLD as an outcome. Nonetheless, it warrants the development of more powerful NAFLD GWAS. A second limitation is that NAFLD was not associated with T2D and CAD in univariable analyses. Although this might contradict the results of observational analyses (Fabbrini et al., 2009; Kotronen & Yki-Järvinen, 2008; Ndumele et al., 2011), caution is needed as some SNPs that were included in these analyses are associated with NAFLD but also with lower LDL cholesterol levels. However, this type of pleiotropy does not influence our observation that genetically predicted waist circumference is associated with T2D and CAD independently of NAFLD.

In conclusion, results of this MVMR investigation suggest that independently of BMI, waist circumference is a strong and causal contributor to NAFLD. Also, the association between waist circumference and T2D and CAD is independent of NAFLD. Altogether, the results put forth that subcutaneous adipose tissue dysfunction and visceral fat accumulation may represent a root cause of a broad range of cardiometabolic diseases. Interventions targeting ectopic lipid deposition may be the key to the treatment of cardiometabolic diseases such as NAFLD, CAD and T2D.

## Methods

### Study populations

We combined information from publicly accessible GWAS summary statistics of European ancestry in a two-sample MR setting. **BMI and waist circumference**: The summary statistics for BMI and waist circumference were obtained from the UK Biobank from 461,460 and 462,166 individuals respectively. Measures from the GIANT consortium were also included. Summary statistics for BMI were obtained from a meta-analysis of up to 125 GWAS for 339,224 European individuals (Locke et al., 2015). Summary statistics for WC were obtained from a meta-analysis of 232,101 individuals (Shungin et al., 2015). **WHR adjustedfor BMI**. WHR adjustedfor BMI was calculated as the ratio of waist and hip circumferences adjusted for BMI in 210,088 individuals from the GIANT consortium (Shungin et al., 2015). **Non-alcoholic fatty liver disease:** Genetic association estimates for a clinical diagnosis of NAFLD were obtained from a recently preprinted GWAS (8434 cases and 770,180 controls) of European ancestry from four cohorts (Ghodsian, 2021). Briefly, we performed a fixed effect GWAS meta-analysis of The Electronic Medical Records and Genomics (eMERGE) (Namjou et al., 2019) network, the UK Biobank, the Estonian Biobank and FinnGen using the *METAL* package (Willer, Li, & Abecasis, 2010). NAFLD was defined by the use of electronic health record codes or hospital records. Logistic regression analysis was performed with adjustment for age,sex, BMI, genotyping site and the first three ancestries based principal components. **Coronary artery disease**. GWAS summary statistics for CAD were obtained from a GWAS on 122,733 cases and 424,528 controls from CARDIoGRAMplusC4D and UK Biobank (van der Harst & Verweij, 2018). Samples from CARDIoGRAMplusC4D were drawn from a mixed population (Europeans, East Asian, South Asian, Hispanic and African American), with the majority (77%) of the participants from European ancestry. Case status was defined by CAD diagnosis, including myocardial infarction, acute coronary syndrome,chronic stable angina or coronary stenosis. **Type 2 diabetes**. GWAS summary statistics for type 2 diabetes were obtained from the DIAbetes Genetics Replication and Meta-analysis (DIAGRAM) consortium and UK Biobank (74,124 cases/824,006 controls) (Mahajan et al., 2018). Case status was defined by a clinical diagnostic of T2D.

### Selection of genetic variants and variants harmonization

For univariable MR analysis, we selected all genome-wide significant SNPs (p-value <5e-8). We then ensured the independence of genetic instruments by clumping all neighbouring SNPs in a 10 Mb window with a linkage disequilibrium R2<0.001 using the European 1000-genome LD reference panel. For multivariable MR analyses, we first extracted all genetic instruments that were previously selected for univariable MR analysis. We then pooled these SNPs to the lowest p-value corresponding to any of the exposures, using the same parameter setting as the univariable MR (R2=0.001 window=10 Mb). When NAFLD was used as an exposure in MVMR, we pooled the combined list of SNPs by selecting the SNP with the lowest p-value for NALFD. This procedure was implemented to select a maximum number of strong genetic instrument, as fewer genetic instruments are available for NAFLD exposure. SNPs in a 2 Mb window of the *HLA, ABO* and *APOE* genetic regions were excluded due to their complex genetic architecture. Harmonization was performed by aligning the effect sizes of different studies on the same effect allele. Palindromic SNPs with a minor allele frequency > 0.3 were removed, otherwise we inferred positive strand alleles with effect allele frequencies. When a particular SNPwas not present in the outcome datasets, we used a proxy SNPs (R2> 0.8). We used the *LDlinkR* V.1.1.2 package (Myers, Chanock, & Machiela, 2020) to interrogate the LDlink API (Machiela & Chanock, 2015) and obtain linkage disequilibrium matrix of European samples from the 1000 Genomes Project.

### Statistical analyses

For univariable primary MR analysis, we performed the inverse-variance weighted (IVW) method with multiplicative random effects with a standard error correction for under dispersion (Burgess, Foley, & Zuber, 2018). MR estimates bias occurs if the genetic instruments influence several traits on different causal pathways. This phenomenon, referred to as horizontal pleiotropy, can be balanced by using multiple genetic variants combined with robust MR methods (Slob & Burgess, 2020). To verify if pleiotropy likely influenced the primary MR results, we performed 5 different robust MR analyses: the MR-Robust Adjusted Profile Score (MR-RAPS) (Zhao, Wang, Bowden, & Small, 2018), the contamination mixture (Slob & Burgess, 2020), the weighted median, the weighted mode and the MR-PRESSO (Verbanck, Chen, Neale, & Do, 2018), each making a different assumption about the underlying nature of the pleiotropy. Consistent estimates across methods provides further confirmation about the nature of the causal links. All continuous exposure estimates were normalized and reported on a standard deviation scale. For dichotomous traits (i.e., diseased status GWAS on NAFLD, T2D and CAD), odds ratios were reported. For multivariable MR analysis, we conducted the IVW method (Gormley et al., 2020). The use of multivariable MR is analogous to the inclusion of measured covariates in a multivariate linear regression. Multivariable MR uses a set of overlapping genetic instrument to estimate the direct effect of an exposure on an outcome. Multivariable MR-IVW analyses were performed using the *MendelianRandomization* V.0.5.1 package (Yavorska & Burgess, 2017).

### Data and code availability

GWAS summary statistics for anthropometric traits from GIANT are available at: https://portals.broadinstitute.org/collaboration/giant/index.php/GIANT_consortium_data_files GWAS summary statistics for BMI from UKB are available via the MR Base GWAS catalogue at id “ukb-b-19953”.

GWAS summary statistics for WC from UKB are available via the MR Base GWAS catalogue at id “ukb-b-9405”.

GWAS summary statistics for T2D are available at: http://diagramconsortium.org/downloads.html

GWAS summary statistics for CAD are available at: https://www.cardiomics.net/download-data

The *TwoSampleMR* package is available at: https://github.com/MRCIEU/TwoSampleMR

The *MendelianRandomization* package is available at: https://github.com/cran/MendelianRandomization

The *data*.*table* package is available at https://github.com/Rdatatable/data.table

The *tidyverse* package collection is available at: https://github.com/tidyverse/tidyverse

The *LDlinkR* package is available at: https://github.com/CBIIT/LDlinkR.

## Data Availability

GWAS summary statistics for anthropometric traits from GIANT are available at: https://portals.broadinstitute.org/collaboration/giant/index.php/GIANT_consortium_data_files GWAS summary statistics for BMI from UKB are available via the MR Base GWAS catalogue at id "ukb-b-19953". GWAS summary statistics for WC from UKB are available via the MR Base GWAS catalogue at id "ukb-b-9405". GWAS summary statistics for T2D are available at: http://diagramconsortium.org/downloads.html GWAS summary statistics for CAD are available at: https://www.cardiomics.net/download-data The TwoSampleMR package is available at: https://github.com/MRCIEU/TwoSampleMR The MendelianRandomization package is available at: https://github.com/cran/MendelianRandomization The data.table package is available at https://github.com/Rdatatable/data.table The tidyverse package collection is available at: https://github.com/tidyverse/tidyverse The LDlinkR package is available at: https://github.com/CBIIT/LDlinkR.

## Acknowledgements

We would like to thank all study participants as well as all investigators of the studies that were used throughout the course of this investigation. WP holds a masters research award from the Canadian Institutes of Health Research (CIHR). EG and IMP hold a doctoral research award from the *Fonds de recherche du Québec: Santé*. (FRQS). BJA and ST hold junior scholar awards from the FRQS. MCVis Canada Research Chair in Genomics applied to Nutrition and Metabolic Health. AT is co-Director of the Research Chair in Bariatric and Metabolic Surgery at Laval University. Part of this study was supported by the European Union through the European Regional Development fund. The work of Estonian Genome Center, Univ. of Tartu has been supported by the European Regional Development Fund and grants No. GENTRANSMED (2014-2020.4.01.15-0012), MOBERA5 (Norface Network project no 462.16.107) and 2014-2020.4.01.16-0125. This study was also funded by the European Union through Horizon 2020 research and innovation programme under grant no 810,645 and through the European Regional Development Fund.

