## Supplementary Figures for "A multivariable Mendelian randomization analysis disentangling the causal relations between abdominal obesity, non-alcoholic fatty liver disease and cardiometabolic diseases"

**Supplementary Materials**

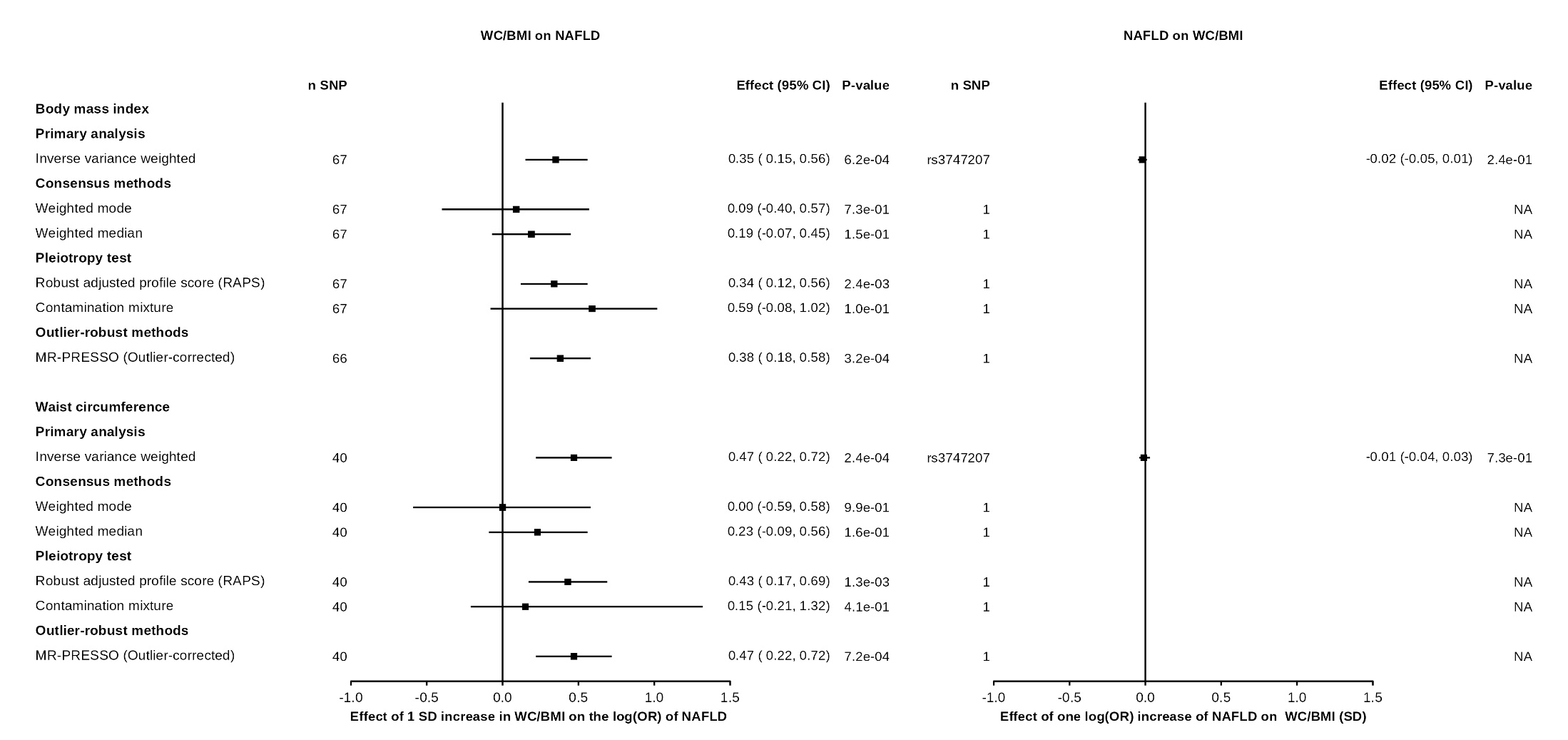

**Supplementary Figure 1. Bi-directional associations between obesity from GIANT and non-alcoholic fatty liver disease (NAFLD).** A) association between obesity and body fat distribution indices (exposures) and NAFLD (outcome) using inverse-variance weighted Mendelian randomization (IVW-MR) and all sensitivity analysis B) association NAFLD (exposure) and obesity and body fat distribution indices (outcomes) using IVW-MR.

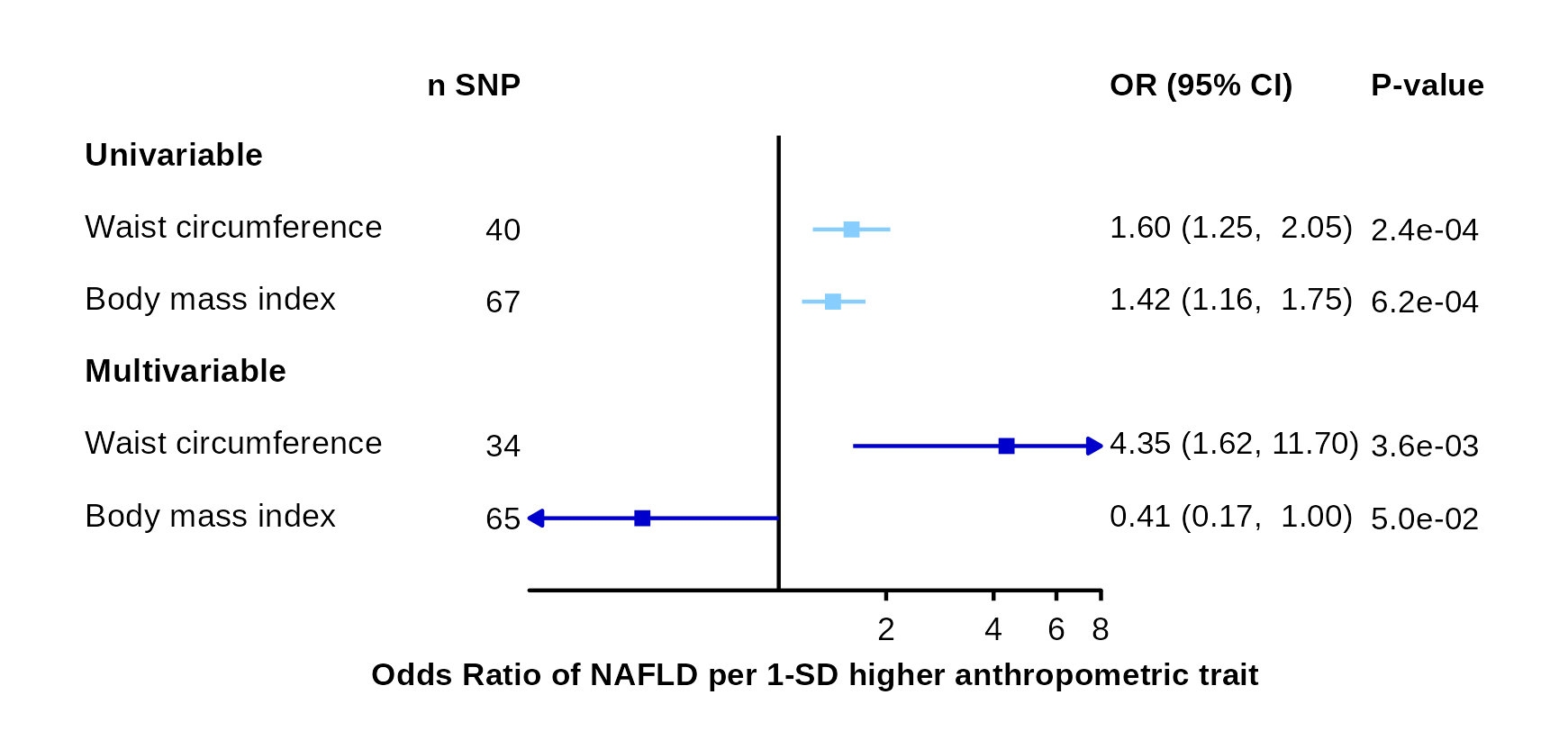

### Supplementary Figure 2. Association between waist circumference and body mass index from GIANT with non-alcoholic fatty liver disease using multivariable Mendelian randomization.

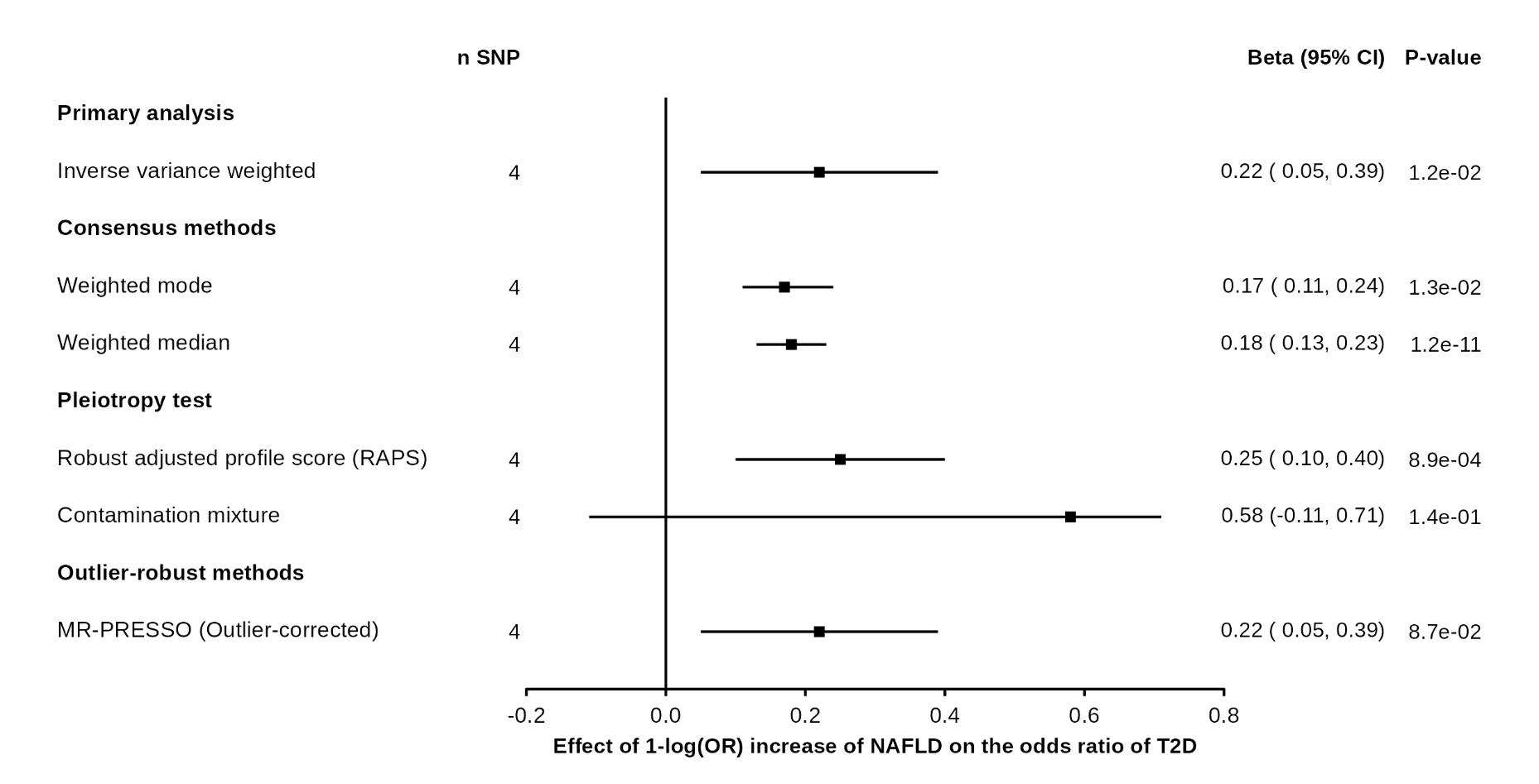

**Supplementary Figure 3. Association between non-alcoholic fatty liver and type 2 diabetes with various MR methods.**

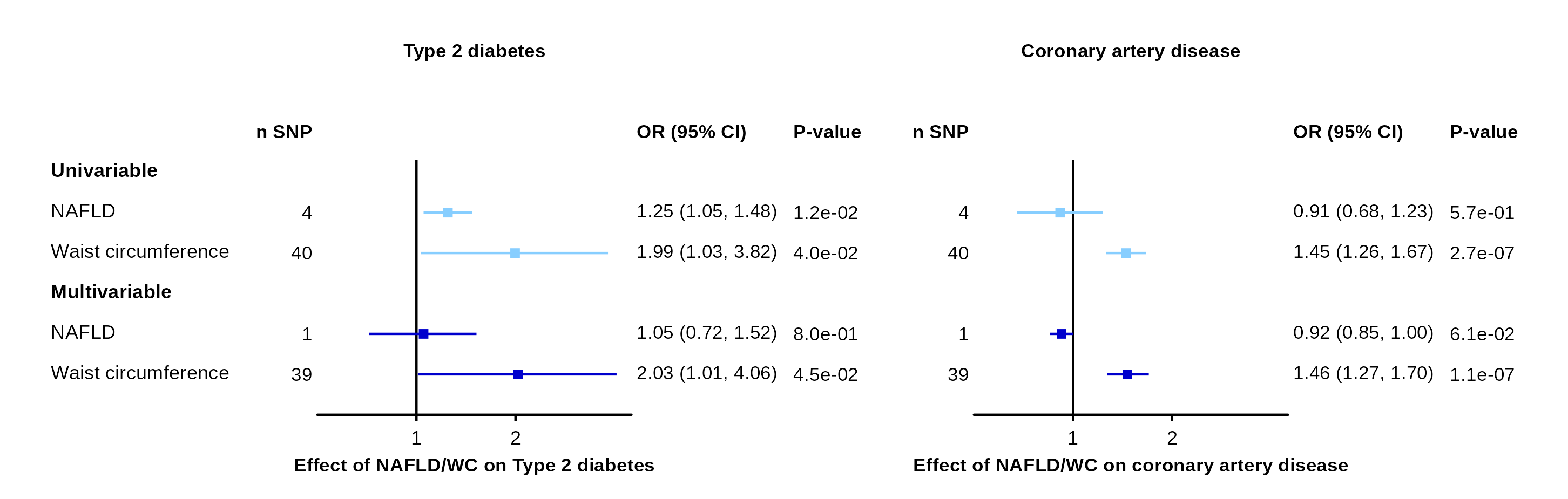

**Supplementary Figure 4. Association between waist circumference from GIANT and coronary artery disease and type 2 diabetes before and after accounting for non-alcoholic fatty liver disease using univariable and multivariable Mendelian randomization.**
